# The impact of COVID-19 infection on social care use in people over 50 years of age: a matched cohort study

**DOI:** 10.1101/2025.02.03.25321574

**Authors:** Rebecca Butfield, Benjamin D Bray, Jennifer Quint, Alejandra Castanon, Elizabeth Adesanya, Simon Chen, Rachel Russell

## Abstract

**Objectives:** To estimate the impact of COVID-19 infection on the requirement for social care services among adults aged ≥50 years in North-West London.

**Design:** Population based matched cohort study using linked routinely collected electronic social care, primary care and hospital records (the Discover dataset).

**Setting:** Approximately 4.7 million people with a general practitioner record in North-West London.

**Participants:** 150,654 adults aged ≥50 years with a first diagnosis of COVID-19 between January 2020 and February 2023 and 547,704 propensity score matched comparators without a COVID-19 diagnosis during the same period.

**Main outcome measures:** Social care use and associated costs overall and by specific type (care home, domiciliary care, respite care, social care assessments) stratified by age group, index year, diagnosis setting, severe COVID-19 risk status, frailty, and care home admission prior to index. Overall survival.

**Results:** A total of 9,174 (6.09%) individuals with COVID-19 required social care use (of any type) during follow up, 2.54 times (95%CI: 2.48-2.61; P<0.0001) higher than matched comparators (n=13,126, 2.40%). The difference was largest for care home admission; individuals with COVID-19 had 4.10 (95%CI: 3.87-4.36; P<0.0001) times the risk of a care home admission and nearly twice the risk (risk ratio 1.94; 95%CI:1.86-2.02 P<0.0001) of domiciliary care during follow up compared with matched comparators. Individuals with COVID-19 experienced higher mortality, with 9.30% (14,005/547,704) dying during follow up compared with 3.76% (20,608/547,704) deaths among matched comparators.

This increase in social care utilisation was observed for all age groups. Adults with COVID-19 had over four times higher social care costs than matched comparators (£1,276 per person per year (pppy) versus £276 pppy; mean difference +£1000, 95%CI: £947-£1054, p<0.0001), with most of the costs due to care home admissions. Higher social care costs in individuals with COVID-19 compared with matched comparators were strongly age related, rising from a mean difference of £130 pppy (95%CI: £99-£161) in those aged 50-64 to £6108 pppy (95%CI: £5613-£6603) higher costs in those aged ≥ 85.

**Conclusions:** COVID-19 infection is associated with meaningfully higher social care requirements in the ≥50 years population. Reducing the need for social care use and the associated costs of care should be one of the goals of interventions to reduce the risk and severity of COVID-19 infection.

## Introduction

Coronavirus disease 2019 (COVID-19), caused by the severe acute respiratory syndrome-coronavirus-2 (SARS-CoV-2), is a highly infectious respiratory illness associated with considerable clinical consequences including hospitalisation or intensive care admission^1^. The COVID-19 pandemic caused major disruptions to healthcare, economic, and social systems worldwide. In the United Kingdom, the pandemic resulted in approximately 25 million cases and over 232,112 deaths as of September 2024^2,3^. Risk factors such as older age, a weakened immune system, pre-existing comorbidities, obesity, smoking, and being from a minority ethnic group increase the morbidity and severity in COVID-19^4,5^. The clinical and economic burden of COVID-19 in the healthcare setting is well established. For example, in 2020, direct healthcare costs for managing COVID-19 in the UK were estimated at £40 billion^6^, while the broader economic impact of government measures ranged between £310 billion and £410 billion, equivalent to £4,600 to £6,100 per person^7^. A recent study quantifying the healthcare resource utilisation and costs among adults with COVID-19 reported that median healthcare costs per hospitalisation was higher in those aged 75–84 (£8942) and ≥85 years (£8835) than in those aged <50 years (£7703)^8^.

Despite previous research reporting the healthcare resource utilisation and costs among adults with COVID-19, there is a lack of evidence on subsequent burden in the social care setting. In the United Kingdom, social care refers to care provided due to age, illness, disability, or other circumstances^9^. Social care can include care administered in the community (e.g., community nursing), residential (e.g., care homes and nursing homes), or domiciliary (e.g., care at home and support workers) settings, along with reablement services that aim to maximise independence and reduce the need for long-term health or social care^10,11^. Insufficient social care capacity has been identified as a key factor in delayed hospital discharges, underscoring the critical link between healthcare and social care in managing patient flow and reducing hospital bed pressures^12^.

To our knowledge, no studies have reported the social care use associated with COVID-19 infection. We therefore aimed to generate evidence to quantify the impact of COVID-19 infection on social care use in adults in North-West London. By addressing this evidence gap, we will provide valuable evidence of the burden of disease in the social care system, allowing a system-wide approach to cost-effectiveness analysis which would facilitate better-informed policy decisions.

## Methods

### Data source and study design

We conducted a matched cohort study using linked primary, secondary and social care real-world data from the Discover database between 1st January 2015 and 29th February 2024 to compare individuals with a COVID-19 diagnosis to propensity score matched comparators without COVID-19 (study design schematic shown in **Appendix Figure S1**). The study is reported using the STROBE-RECORD guidelines (**Appendix S1**).

Discover is a research dataset of health and social care records for a population of >4.7 million residents of North-West London (NWL) who are registered with a general practitioner (GP). Discover is broadly representative of the UK population by age and sex, but is more diverse in terms of ethnicity and deprivation^13^.

### Study population

Adults aged ≥50 years registered with a GP in NWL who had: (1) no missing age, sex, ethnicity, or index of multiple deprivation (IMD) data; (2) data considered of acceptable research quality during the study period (e.g., excluding patients with poor data recording that raises suspicion as to the validity of that patient’s record); and (3) at least 12 months follow up available prior to the end of the study period or exit from the Discover database were eligible for inclusion (**Figure 1**). Adults with a date of death in the first 12 months of follow up were eligible for inclusion into the study.

**Figure 1:**
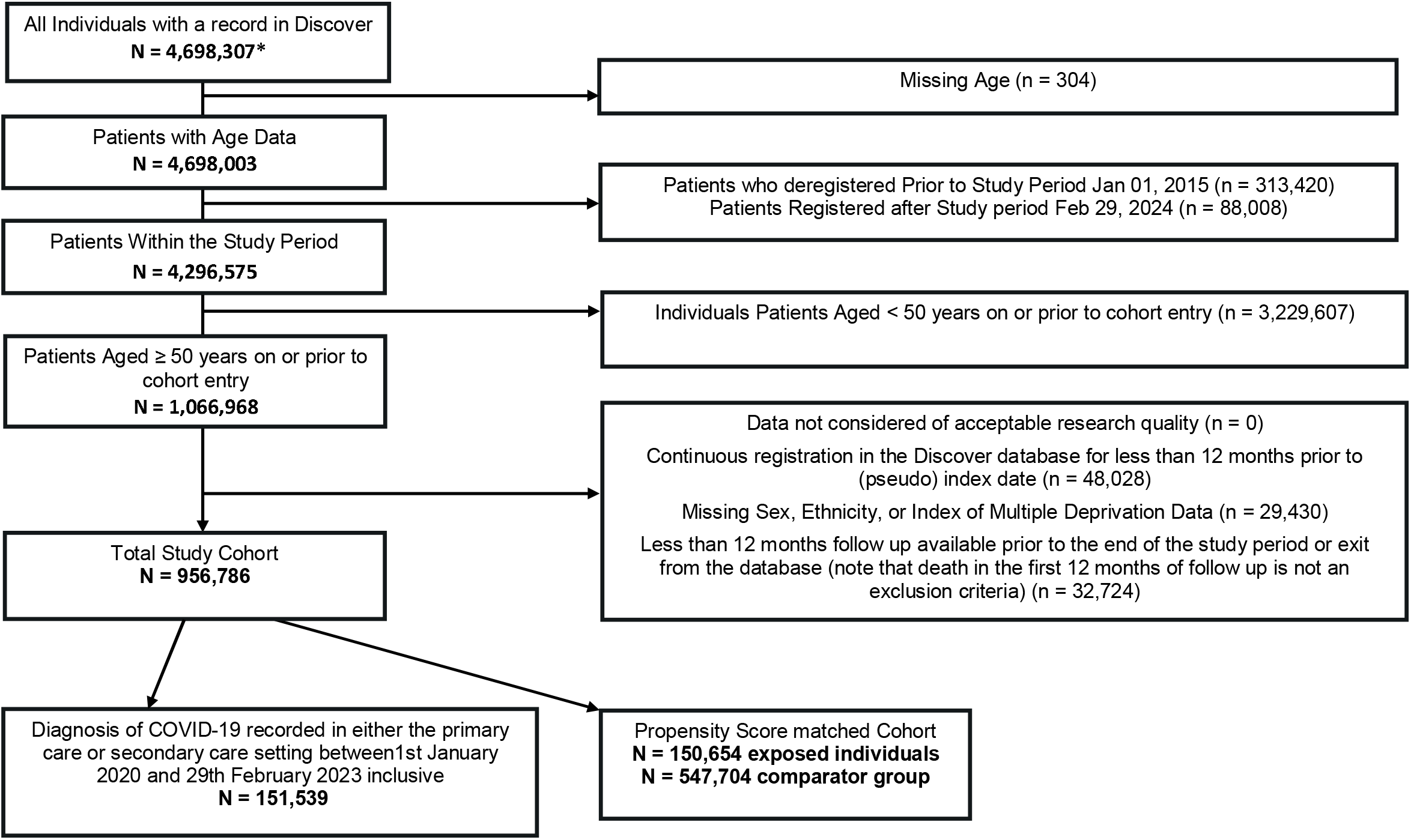
Flow diagram showing the creation of the cohort and reasons for exclusion

Adults with a first diagnosis of COVID-19 (cases) recorded in either primary or secondary care between 1st January 2020, and 28th February 2023 (indexing period) were propensity score matched to up to four adults without a diagnosis of COVID-19 at the time their matched case was diagnosed. We followed individuals from the date of their COVID-19 diagnosis (index date). Comparators were assigned the same date as their matched case with COVID-19 (pseudo index date). Follow up ended at the earliest of study end (29^th^ February 2024), the date of transfer out of the NWL region, or death. Comparators with a diagnosis of COVID-19 during the indexing period were censored from the comparator group and became cases, while those with a diagnosis after the end of the indexing period remained in the comparator group until the end of the study.

### Exposures

COVID-19 diagnosis was ascertained using diagnostic Read (version 2) or SNOMED codes recorded in primary care, or International Classification of Diseases, Tenth Revision (ICD-10) codes recorded in secondary care inpatient admissions. Code lists were adapted by the study team from previously published research identifying COVID-19 diagnosis in electronic health records in England^14^. The adapted code lists were reviewed by multiple reviewers and verified by those with clinical experience within the UK health service and deemed appropriate for use (**Appendix S2**).

### Propensity score matching

Propensity scores were based on the probability of being diagnosed with COVID-19 during the indexing period given characteristics at the time of meeting cohort eligibility. Scores were estimated by fitting a logistic regression model with age in years, sex, ethnicity, deprivation deciles, body mass index (BMI), smoking status, and severe COVID-19 risk status included as variables. Covariate distribution balance pre- and post-matching was checked using summary statistics, absolute standardised mean differences (aSMD, with aSMD <0.1 indicating good balance^15^) and density plots visualising propensity score distribution overlap (**Appendix S3**).

### Outcomes

We identified social care resource use (SCRU) recorded in the administrative ‘Service Type Group’ and ‘Service Type Description’ fields in the electronic social care records. Automated code was used to search for keywords identifying specific types of social care use, as per a previous study using electronic social care records in Discover^16^. We classified social care resource use into four pre-defined categories – care home, domiciliary care, respite care, and social care assessment. SCRU and associated costs were estimated overall and for each social care category. We transformed SCRU into direct costs using unit cost data sourced the Unit Costs of Health and Social Care 2023^17^.

The following unit costs were used: (1) £247 per day for care home; (2) £27 per hour for domiciliary care with an assumption of 7.4 hours of care provided per day; (3) £68 per day for respite care; and (4) £49 per social care assessment. A summary of the operational definitions of social care use and costs are described in **Appendix S4**.

### Demographic and clinical variables

Demographic characteristics included age, sex, ethnicity, BMI, smoking status, deprivation (measured using deciles of index of multiple deprivation), and geolocation of GP practice. BMI and smoking status were identified using the most recent data within two years either side of the individuals (pseudo) index date, due to anticipated infrequency in reporting^18^. Clinical characteristics included admission to care home prior to index, frailty (derived from both electronic and clinical frailty indices, **Appendix S5**), and at high risk of severe COVID-19. Severe COVID-19 risk status was defined using the clinical risk groups outlined in COVID-19: the Green Book, Chapter 14a.^19^ Individuals identified with presence of any of the comorbidities reported in the Green Book were classified ‘High-risk of severe COVID-19’, while those without were categorised ‘Not at high-risk of severe COVID-19’. Further details on definitions and operationalisation of code lists are described in **Appendix S5**.

### Statistical analyses

We described the demographic and clinical characteristics of COVID-19 cases and matched comparators at (pseudo) index date. Numbers and percentages were used for categorical data and mean (standard deviation, SD) or median (interquartile range, IQR) were used for continuous data. Kaplan-Meier curves were used to plot survival.

We reported the number and proportion of individuals with at least one social care record (event) of any type and of each pre-defined social care category (care home, domiciliary care, respite care, social care assessment) during follow up. Risk ratios and 95% confidence intervals (CIs) were calculated to compare the proportions with at least one SCRU record between adults with COVID-19 and matched comparator cohort. Chi-squared tests were used to estimate the strength of evidence for a difference in proportions for social care resource use.

Mean (SD) social care events recorded per person per year (pppy), length of stay in care home, respite care, and domiciliary care pppy, and social care costs pppy were reported overall, and by each social care type. Mean differences and associated 95% CIs were estimated for all three outcomes. Rate ratios and 95% CIs were used to assess the event rate of SCRU between adults with COVID-19 and the comparator cohort. Length of stay for adults in respite or domiciliary care was calculated from the start date of social care until the earliest of the end date of social care, date of care home admission after the start date of social care, or the end of study date. Adults in care homes were considered resident from the date of admission for the duration of follow up. Wilcoxon signed-rank tests were used to estimate the strength of evidence for a difference in means for social care events, length of stay and costs.

Outcomes were evaluated by age group, index year, diagnosis setting, severe COVID-19 risk status, frailty, and care home admission prior to index. We performed a secondary analysis comparing social care use 12-months before and after COVID-19 infection to explore SCRU among all eligible individuals with COVID-19 including those not taken forward to the matched cohort (**Appendix S6**). We repeated our main analyses in four sensitivity analyses to assess the robustness of our findings (**Appendix S7**). All analyses were conducted according to a statistical analysis plan in R (Version 3.6.1).

### Results

Of a total population of 4,698,307 individuals, 956,786 were eligible for study inclusion. A total of 151,539 adults with COVID-19 were identified. The propensity score matching process was successful for 150,654 individuals with COVID-19, who were matched to 547,704 comparators (**Figure 1**).

### Demographic and clinical characteristics

The mean age of individuals with COVID-19 was 64 years (SD=11), while the mean age of matched comparators was 63 years (SD=11). Individuals with COVID-19 and matched comparators were broadly similar in age, sex, ethnicity, and deprivation at (pseudo) index (**Table 1**). Compared with matched comparators, a higher frequency of individuals with COVID-19 had: a record of frailty (28.07% vs 22.51%), were categorised as overweight, obese, or severely obese according to BMI (54.05% vs 48.82%) and were at high risk of severe COVID-19 (13.53% vs 11.91%). Of adults with known smoking status, 27.14% of individuals with COVID-19 were current or previous smokers, compared with 24.93% of matched comparators. Individuals with COVID-19 were also more likely to be admitted to a care home prior to index compared with matched comparators (0.66% and 0.11% respectively).

**Table 1:** Demographic and clinical characteristics of COVID-19 cases and matched comparators at index. Values are numbers (corresponding percentages) unless otherwise stated.

| Variables | Matched COVID-19 cases<br>(n = 150,654) | Matched comparators<br>(n = 547, 704) |
| --- | --- | --- |
| **Age, years** |  |  |
| Mean (Standard Deviation) | 64 (11) | 63 (11) |
| Median (Interquartile Range) | 61 (55-71) | 60 (53-70) |
| **Age group, years** |  |  |
| 50-64 | 91,250 (60.57) | 340,146 (62.10) |
| 65-74 | 31,057 (20.61) | 114,009 (20.82) |
| 75-84 | 18,361 (12.19) | 64,784 (11.83) |
| ≥85 | 9,986 (6.63) | 28,765 (5.25) |
| **Sex** |  |  |
| Male | 71,173 (47.24) | 259,084 (47.30) |
| Female | 79,481 (52.76) | 288,620 (52.70) |
| **Ethnicity** |  |  |
| White | 80,132 (53.19) | 283,474 (51.76) |
| Asian or Asian British | 43,020 (28.56) | 158,131 (28.87) |
| Black or Black British | 13,053 (8.66) | 49,962 (9.12) |
| Mixed | 3,642 (2.42) | 13,936 (2.54) |
| Other | 10,807 (7.17) | 42,201 (7.71) |
| **BMI** |  |  |
| Underweight (<18.5 kg/m<sup>2</sup>) | 2,865 (1.90) | 7,701 (1.41) |
| Healthy (18.5-24.9 kg/m<sup>2</sup>) | 37,585 (24.95) | 123,938 (22.63) |
| Overweight (25-29.9 kg/m<sup>2</sup>) | 44,281 (29.39) | 147,784 (26.98) |
| Obese (30-39.9 kg/m<sup>2</sup>) | 32,281 (21.43) | 104,930 (19.16) |
| Severely Obese (≥40 kg/m<sup>2</sup>) | 4,870 (3.23) | 14,701 (2.68) |
| Unknown | 28,772 (19.10) | 148,650 (27.14) |
| **IMD decile** |  |  |
| 1 (most deprived) | 3,167 (2.10) | 11,752 (2.15) |
| 2 | 14,361 (9.53) | 53,409 (9.75) |
| 3 | 20,585 (13.66) | 75,112 (13.71) |
| 4 | 23,083 (15.32) | 85,467 (15.60) |
| 5 | 22,257 (14.77) | 81,250 (14.83) |
| 6 | 20,356 (13.51) | 73,811 (13.48) |
| 7 | 15,253 (10.12) | 54,620 (9.97) |
| 8 | 14,662 (9.73) | 53,405 (9.75) |
| 9 | 11,219 (7.45) | 39,665 (7.24) |
| 10 (least deprived) | 5,711 (3.79) | 19,213 (3.51) |
| **Smoking status** |  |  |
| Current smoker | 11,658 (7.74) | 50,766 (9.27) |
| Previous smoker | 29,222 (19.40) | 85,757 (15.66) |
| Never smoked | 81,795 (54.29) | 268,653 (49.05) |
| Missing | 27,979 (18.57) | 142,528 (26.02) |
| **Frailty** |  |  |
| Mild | 14,468 (9.60) | 51,333 (9.37) |
| Moderate | 17,033 (11.31) | 51,046 (9.32) |
| Severe | 9,732 (6.46) | 18,530 (3.38) |
| Frail with unspecified severity | 1,056 (0.70) | 2,383 (0.44) |
| Unknown | 108,365 (71.93) | 424,412 (77.49) |
| **Geolocation** |  |  |
| Central London (Westminster) | 11,044 (7.33) | 53,162 (9.71) |
| West London (Kensington and Chelsea) | 8,856 (5.88) | 42,427 (7.75) |
| Brent | 23,270 (15.45) | 80,672 (14.73) |
| Ealing | 24,227 (16.08) | 89,050 (16.26) |
| Hammersmith and Fulham | 10,547 (7.00) | 43,782 (7.99) |
| Harrow | 21,521 (14.29) | 65,659 (11.99) |
| Hillingdon | 25,425 (16.88) | 76,964 (14.05) |
| Hounslow | 17,990 (11.94) | 85,810 (12.02) |
| Other | 7,774 (5.16) | 30,178 (5.51) |
| **Risk of severe COVID-19** |  |  |
| High-risk of severe COVID | 20,384 (13.53) | 65,219 (11.91) |
| Not at high-risk of severe COVID | 130,270 (86.47) | 482,485 (88.09) |
| **Admission to care home prior to index** |  |  |
| Admission to Care Home Prior to Index | 988 (0.66) | 600 (0.11) |
| No Admission to Care Home Prior to Index | 149,666 (99.34) | 547,104 (99.89) |

**Abbreviations:** BMI, Body Mass Index; IMD, Index of Multiple Deprivation

### Main analysis

A total of 9,174 (6.09%) of individuals with COVID-19 required social care of any type during follow up, 2.54 times (95%CI: 2.48-2.61; p<0.0001) higher than the matched comparators (n=13,126, 2.40%) (**Table 2**). The difference was largest for care home admission, individuals with COVID-19 having 4.10 (95%CI: 3.87-4.36; p<0.0001) times the risk of admission to a care home during follow up compared with matched comparators. Individuals with COVID-19 were almost twice as likely to require domiciliary care (Risk ratio 1.94; 95%CI: 1.86-2.02 p<0.0001) than matched comparators.

**Table 2:** Number, proportion, and associated risk ratios of COVID-19 cases and matched comparators with at least one social care record during follow up.

|  | **COVID-19 cases**<br>**N (%)** | **Matched comparators**<br>**N (%)** | **P-value\*** | **Risk ratio (95% CI)** |
| --- | --- | --- | --- | --- |
| **Individuals with at least one social care record** | 9,174 (6.09) | 13,126 (2.40) | <0.0001 | 2.54 (2.48, 2.61) |
| **Individuals with at least one care home admission** | 2,288 (1.52) | 2,027 (0.37) | <0.0001 | 4.10 (3.87, 4.36) |
| **Individuals with at least one domiciliary care record** | 3,365 (2.23) | 6,314 (1.15) | <0.0001 | 1.94 (1.86, 2.02) |
| **Individuals with at least one respite care record** | 51 (0.03) | 88 (0.02) | <0.0001 | 2.11 (1.49, 2.97) |
| **Individuals with at least one social care assessment record** | 4,871 (3.23) | 6,474 (1.18) | <0.0001 | 2.74 (2.64, 2.84) |

\*Chi-squared tests were used to generate p-values estimating the strength of evidence for a difference in proportions for social care records

An increase in the rate of social care events was observed for all age groups but was highest for the ≥ 85 years age group (Rate ratio (RR) for all types of social care 3.86, 95% CI: 3.77-3.96; p<0.0001, versus rate ratios of 1.49, 3.21 and 3.48 for the 50-64 years, 65-74 years and 75-84 years groups respectively) (**Figure 2**). Increased rate of care home admissions in individuals with COVID-19 was particularly high for individuals with a record of frailty (RR 6.23, 95%CI: 5.80-6.70; p<0.0001) but was also observed for individuals without frailty (RR 1.88, 95%CI: 1.66-2.12; p<0.0001). Mean length of stay in domiciliary care and care home was higher among individuals with COVID-19, however length of stay in respite care was higher among matched comparators (**Table 3**).

**Table 3:**
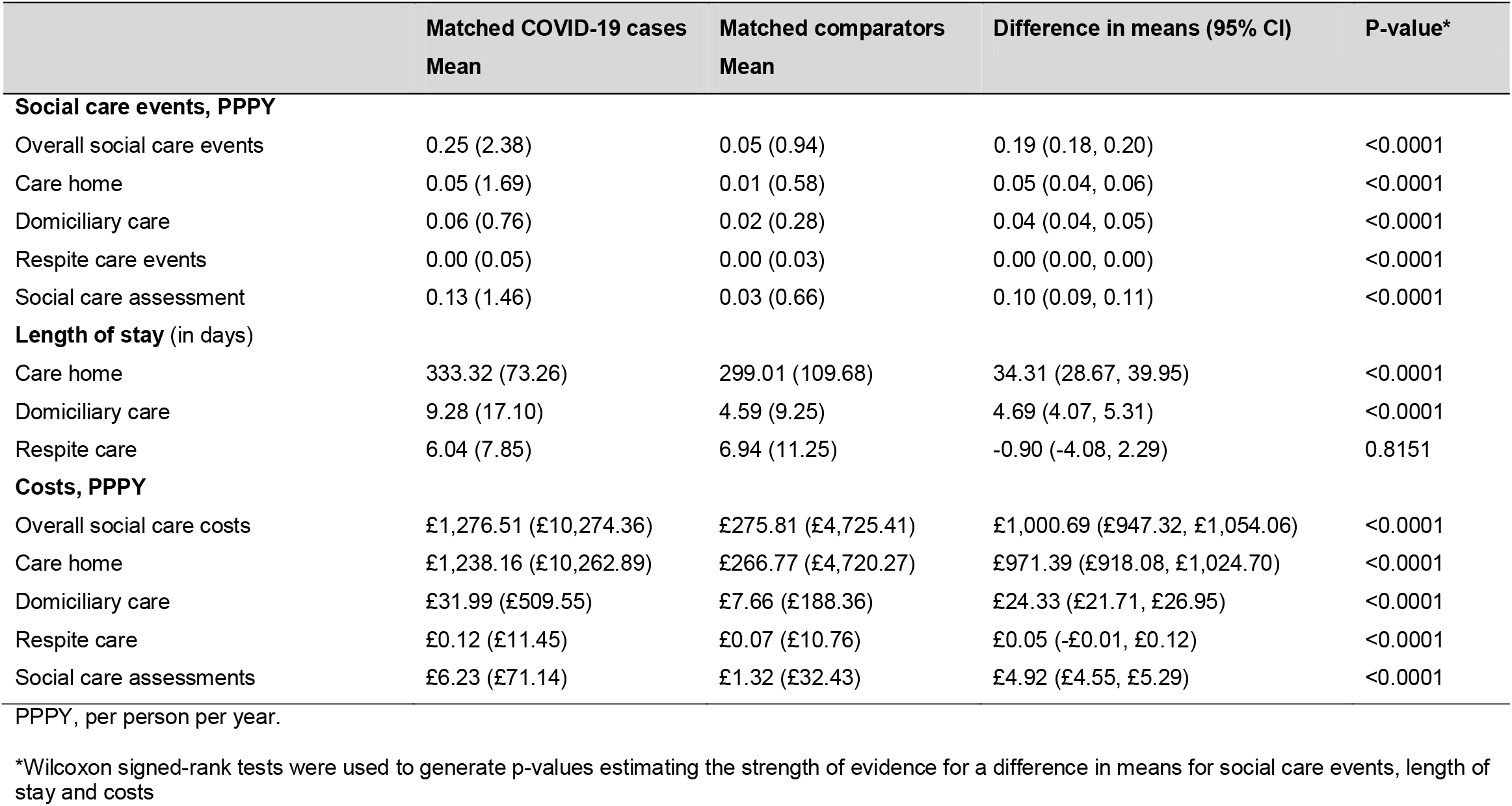
Mean social care events, length of stay and associated costs during follow up. Values are mean (corresponding standard deviation) unless otherwise stated.

|  | Matched COVID-19 cases<br>Mean | Matched comparators<br>Mean | Difference in means (95% CI) | P-value* |
| --- | --- | --- | --- | --- |
| **Social care events, PPPY** |  |  |  |  |
| Overall social care events | 0.25 (2.38) | 0.05 (0.94) | 0.19 (0.18, 0.20) | <0.0001 |
| Care home | 0.05 (1.69) | 0.01 (0.58) | 0.05 (0.04, 0.06) | <0.0001 |
| Domiciliary care | 0.06 (0.76) | 0.02 (0.28) | 0.04 (0.04, 0.05) | <0.0001 |
| Respite care events | 0.00 (0.05) | 0.00 (0.03) | 0.00 (0.00, 0.00) | <0.0001 |
| Social care assessment | 0.13 (1.46) | 0.03 (0.66) | 0.10 (0.09, 0.11) | <0.0001 |
| **Length of stay (in days)** |  |  |  |  |
| Care home | 333.32 (73.26) | 299.01 (109.68) | 34.31 (28.67, 39.95) | <0.0001 |
| Domiciliary care | 9.28 (17.10) | 4.59 (9.25) | 4.69 (4.07, 5.31) | <0.0001 |
| Respite care | 6.04 (7.85) | 6.94 (11.25) | -0.90 (-4.08, 2.29) | 0.8151 |
| **Costs, PPPY** |  |  |  |  |
| Overall social care costs | £1,276.51 (£10,274.36) | £275.81 (£4,725.41) | £1,000.69 (£947.32, £1,054.06) | <0.0001 |
| Care home | £1,238.16 (£10,262.89) | £266.77 (£4,720.27) | £971.39 (£918.08, £1,024.70) | <0.0001 |
| Domiciliary care | £31.99 (£509.55) | £7.66 (£188.36) | £24.33 (£21.71, £26.95) | <0.0001 |
| Respite care | £0.12 (£11.45) | £0.07 (£10.76) | £0.05 (-£0.01, £0.12) | <0.0001 |
| Social care assessments | £6.23 (£71.14) | £1.32 (£32.43) | £4.92 (£4.55, £5.29) | <0.0001 |

PPPY, per person per year.
\*Wilcoxon signed-rank tests were used to generate p-values estimating the strength of evidence for a difference in means for social care events, length of stay and costs

**Figure 2.**
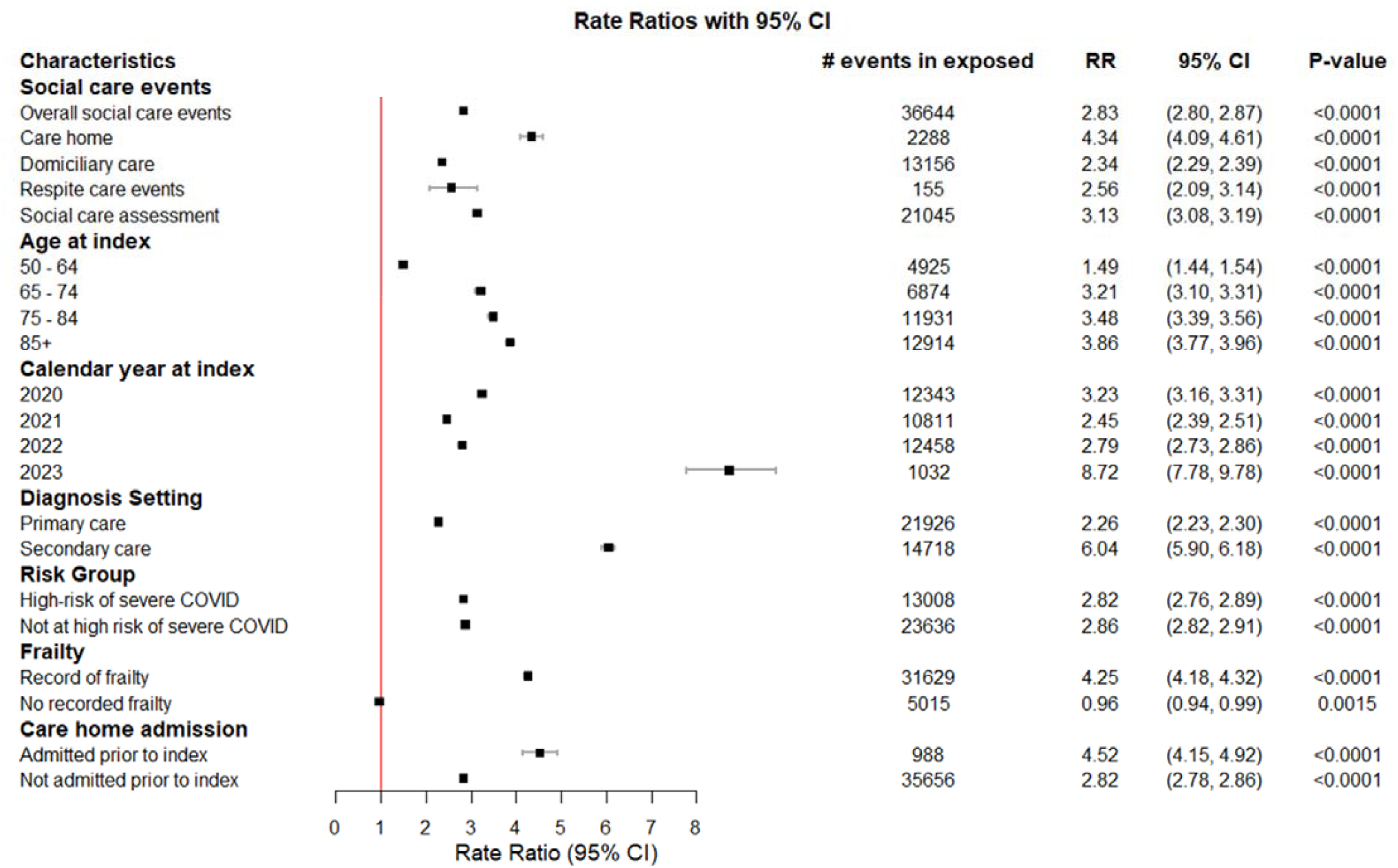
Social care event rate ratios (95% CI) overall and by subgroup comparing people with COVID-19 to propensity-score matched comparators.

Individuals with COVID-19 had over four times higher social care costs than matched comparators (£1,276 pppy versus £276 pppy; mean difference +£1000, 95%CI: £947-£1054, p<0.0001). Most of the costs associated with social care use were due to care home admissions (**Table 3)**, with a mean cost for care home use of £1,238 pppy (SD=£10,263) among individuals with COVID-19, and £267 pppy (SD=£4,720) among matched comparators. Higher social care costs in individuals with COVID-19 compared with comparators were strongly age related, rising from a mean difference of £130 pppy (95%CI: £99-£161) in those aged 50-64 to £6,108 pppy (95%CI: £5,613-£6,603) higher costs in individuals ≥ 85 (**Figure 2**).

Individuals with COVID-19 experienced higher mortality, with (9.30% (14,005/150,654) dying during follow up compared with 3.76% (20,608/547,704; Log rank Kaplan-Meier p<0.0001) deaths among matched comparators. Most of this excess mortality occurred in the first two months after COVID-19 infection (**Appendix Figure S2**).

### Secondary and sensitivity analyses

Among the 151,539 individuals with COVID-19 included in the secondary analysis, the proportions with at least one social care record of any type increased in the 12-months after COVID-19 infection, driven by increases in care home admissions and domiciliary care (**Appendix S6**). Mean costs were £100 pppy higher post COVID-19 than before. Censoring individuals at their second COVID-19 diagnosis yielded similar results to the main analysis (**Appendix S7**). Excluding individuals with social care use prior to index led to a healthier study population and reduced the proportion of COVID-19 cases with at least one social care record to 2.28%. Excluding individuals with care home admission prior to index showed similar results to the main analysis, but of a lower magnitude (**Appendix S7**). Assessing SCRU and costs among individuals with respiratory outcomes in the six months prior to index increased the proportion of individuals with COVID-19 with social care use (19.10% versus 6.09% in the main analysis).

## Discussion

To the best of our knowledge, this is the first study to quantify the impact of COVID-19 infection on social care use and costs in adults aged ≥50 years, using comprehensive linked electronic health and social care records. We identified markedly higher requirements for social care use in individuals with COVID-19 compared with matched comparators, notably for care home admission (approximately four-fold risk) and domiciliary care (approximately double the risk). These increases in social care requirements resulted in over four times higher mean social care costs in individuals with COVID-19. It is plausible that the increase in social care requirements is the result of COVID-19 causing or accelerating physical or cognitive function decline in some survivors, particularly older adults^20,21^.

While evidence on the effect of COVID-19 infection on social care use and associated costs is limited, findings from our study broadly align with other data about social care demands during and after the pandemic. The Association of Directors of Adult Social Services (ADASS) reported an increase in people presenting to local authorities with adult social care needs during the early phases of the pandemic, with a predicted £468m 2020/21 national overspend due to the additional need and costs from the COVID-19 pandemic^22^.

The reasons behind the increased social care burden and costs of COVID-19 observed in this study are likely multifactorial. COVID-19 disproportionately affects elderly and vulnerable populations, with studies suggesting older adults, or those with underlying chronic conditions (e.g., diabetes, cardiovascular disease) are at higher risk of severe infection^4,5^. People recovering from severe COVID-19 infections may experience lasting symptoms and post-COVID syndromes^23^, and individuals with mild or asymptomatic infections can also experience long-term symptoms^24^. These persistent symptoms – termed long-COVID – may require rehabilitation, home care, or other long-term social care services. Notably, we observed increased social care requirements irrespective of individuals’ risk of severe COVID-19 suggesting that even individuals not at higher risk of worse clinical outcomes are at higher risk of social care needs after COVID-19 infection.

Policy changes and non-pharmacological pandemic control measures may have impacted the results of the study. In March 2020, during the initial phase of the COVID-19 pandemic, the UK government published a policy where patients were discharged once clinically safe in order to free up hospital beds^25^. A proportion of patients were discharged to short or long term residential or nursing home care^25^. Changes to legislation led to constraints on care providers, with 70% of respondents from the ADASS survey reporting the largest increase in people presenting increased adult social care needs were people being discharged from hospital^22^. Between September 2020 and March 2021 the government provided £588m to implement the discharge-to-assess model, where social care services were provided and paid for to help people leave hospital before their care needs where assessed^26^.

Our study demonstrates that COVID-19 infection poses substantial pressure and cost to the UK social care system. This study reinforces that policy decisions should consider the economic burden of a disease beyond the healthcare system and the value preventative care, such as the UK COVID-19 vaccination programme, in reducing future social care demand.

### Strengths and limitations

Our study has several limitations. First, the generalisability of results to the wider UK population may be limited as Discover only includes people registered with NWL GPs, and excludes care provided in private medical practice and prisons. However, the Discover database provides near-complete coverage of NWL, reducing selection bias, and is broadly representative of UK population age and sex^13^. Additionally, research by Mendes et al found that Discover is unique in having longitudinal linked health and social care records available for research; until a similar national data source is available, this study addresses a large paucity of evidence on the social care resource use and costs of COVID-19 infections^12^.

Secondly, the identification of social care utilisation in this study may also be limited. There are no standardised or validated definitions for social care resource use in electronic records, therefore we used code to search for keywords. Individuals may therefore be misclassified as having no record of social care during follow up, if the keywords were not present. However, we found that individuals with a social care record of care home admission also had corresponding entries in secondary or primary care records for either admission from or discharge to a care home. Respite care may have been underestimated due to lack of consistency in its recording and it may be conflated with care home admissions. Assumptions regarding the length of social care resource use were implemented due to the absence of distinct social care discharge records. This study reinforces the importance of continuing efforts to develop social care data by the Department of Health and Social Care (DHSC) from the direct perspective of researchers^28,29^, with specific areas of improvement (e.g., standardised definitions, complete recording of social care discharge) highlighted by the data limitations detailed above.

Thirdly, individuals paying for domiciliary care privately or having care provided by lay carers and family members may not have been included in social care records, resulting in underestimation of social care utilisation and costs. Furthermore, the data does not distinguish between privately or publicly funded social care, resulting in difficulties in the identification of costs relating to the Personal Social Services (PSS) – a key distinction required in health technology evaluations by NICE^30^. Despite this, national statistics report that 37% of care home residents between 2022-2023 self-funded their care^31^. Analysis by the Care Quality Commission suggests self-funded admissions can range from 25-51% with an average of 35%, while admissions funded by local authorities have risen to an average of 72%^32^.

COVID-19 was ascertained using diagnosis codes and hence individuals with COVID-19 infection who had not been tested, or who had tested positive but not reported this to healthcare services will be missed. COVID-19 testing practices evolved substantially over the pandemic, including wide use of at-home testing, and increased testing in care homes, however it is difficult to estimate the impact of these changes on the study results. We did not evaluate the effect of vaccination or COVID-19 therapies on social care risk, although this would be a useful question to explore in future studies.

Individuals at high-risk of COVID-19 may not have been identified as certain comorbidities used to identify severe COVID-19 risk status were challenging to identify in this study, either because no standardised definitions were identified in the literature, or some comorbidities (e.g., immune-mediated inflammatory diseases) were defined wholly through prescriptions that are largely issued in secondary care which Discover data does not capture. Where feasible, we aligned comorbidity definitions with previously published studies^8,14^, and verified study specific code lists through clinical input.

## Conclusions

Our study revealed markedly increased social care use and costs associated with COVID-19 infection in adults aged ≥50 years. Interventions to reduce risk and severity of COVID-19 infection could have substantial impact on reducing the need for social care and its associated costs. Our study also highlights the value of improving social care recording in electronic health records to enable accurate estimates of the impact of disease on social care.

## Data Availability

The Discover data that supports the findings of this study is available via Discover-NOW Health Data Research Hub for Real World Evidence through their data scientist specialists and IG committee-approved analysts, hosted by Imperial College Health Partners.

## Other sections requested by journals upon submission

## Acknowledgements

The authors thank Diana Mendes and Carmen Tsang for contributing to the conceptualisation and interpretation of initial findings; Shuk-Li Collings and Kate Mansfield for their interpretation of findings and critical review of this paper.

## Author contributions

RB conceptualised and designed the study, interpreted the data, revised and reviewed the manuscript. RR, JQ, BB, AC designed, interpreted the data, revised and reviewed the manuscript. EA prepared the first draft of the manuscript, contributed to the design, interpreted the data and revised and reviewed the manuscript. SC conducted the analyses, interpreted the data and reviewed the manuscript. BB, AC and SC confirm they had access to and verified the data.

All authors participated in the interpretation of data for the study; critically reviewed and approved the final version of the manuscript and agree to be accountable for all aspects of the work in ensuring that questions related to the accuracy or integrity of any part of the work are appropriately investigated and resolved.

## Ethics statement

This study involved the use of secondary data; no primary data collection was carried out for the purpose of this study. As all patient-level data were fully anonymised, and no direct patient contact or primary collection of individual patient data occurred, patient consent was not required. Discover has secured HRA approval until 2025 to use the Discover Research Platform for research purposes of studies submitted to the NWL Data Access Committee. This study was approved by the NWL Data Access Committee on 16th November 2023, application number ID-375.

## Conflict of interest

RB and RR are employees of Pfizer and may hold stock or stock options. BB is Partner and Head of Health Analytics at Lane Clark & Peacock LLP; AC, EA and SC are employees of Lane Clark & Peacock LLP. JQ reports no conflict of interest.

## Funding

This study was conducted as a collaboration between LCP, JQ and Pfizer. Pfizer is the study sponsor.

EA, AC, SC and BB are employees of Lane Clark & Peacock LLP, which was a paid contractor to Pfizer in connection with the development of this manuscript and were responsible for study management and data analysis.

## Patient and Public involvement

It was not appropriate or possible to involve patients or the public in the design, or conduct, or reporting, or dissemination plans of our research.

