## Appendix for "The impact of COVID-19 infection on social care use in people over 50 years of age: a matched cohort study"

**Supplementary Appendix for the manuscript titled "The impact of COVID-19 infection on social care use in people over 50 years of age: a matched cohort study"**

### Contents


Figure S1: Study design schematic – overview of study design and time periods of interest

Appendix S1: STROBE-RECORD Checklist

Appendix S2: COVID-19 exposure code lists

Appendix S3: Propensity score matching plots

Appendix S4: Social care outcome definitions

Appendix S5: Demographic and clinical characteristic definitions and code lists

Appendix S6: Secondary analysis definition and results

Appendix S7: Sensitivity analyses definitions and results

Figure S2: Main analysis Kaplan-Meier survival plot

**Figure S1 – Overview of the study design and time periods of interest**

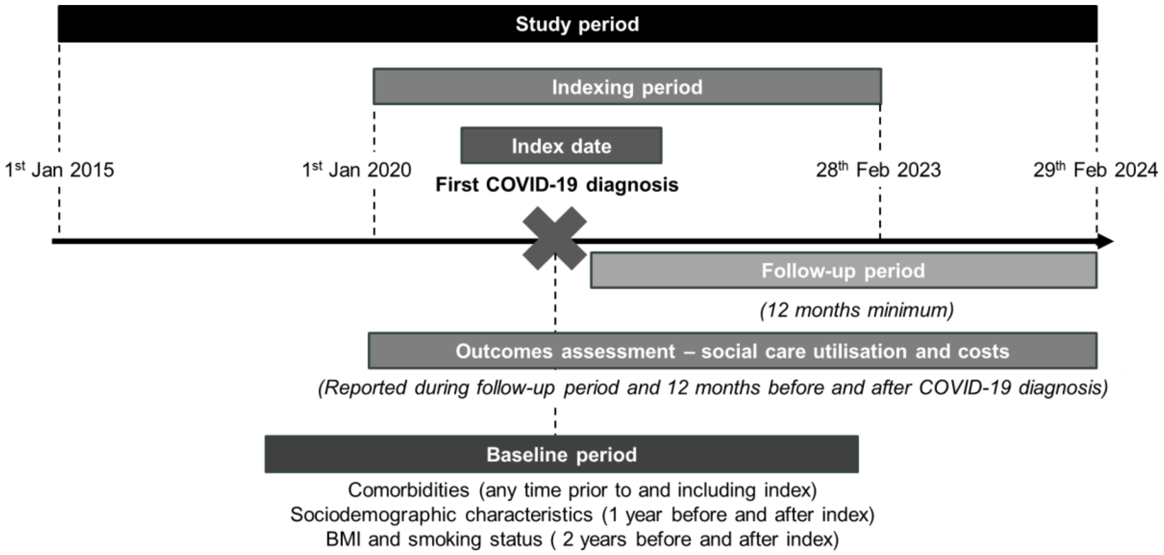

**Appendix S1: STROBE-RECORD checklist**

**The RECORD statement – checklist of items, extended from the STROBE statement, that should be reported in observational studies using routinely collected health data.**

| | **Item No.** | **STROBE items** | **Location in manuscript where items are reported** | **RECORD items** | **Location in manuscript where items are reported** |
|---|---|---|---|---|---|
| **Title and abstract** | | | | | |
| | 1 | (a) Indicate the study's design with a commonly used term in the title or the abstract (b) Provide in the abstract an informative and balanced summary of what was done and what was found | Page 1: Title | RECORD 1.1: The type of data used should be specified in the title or abstract. When possible, the name of the databases used should be included.<br><br>RECORD 1.2: If applicable, the geographic region and timeframe within which the study took place should be reported in the title or abstract.<br><br>RECORD 1.3: If linkage between databases was conducted for the study, this should be clearly stated in the title or abstract. | Page 1: Title<br><br>Page 2: Abstract: Design |
| **Introduction** | | | | | |
| Background rationale | 2 | Explain the scientific background and rationale for the investigation being reported | Page 3: Introduction | | |

| | | | | | |
|---|---|---|---|---|---|
| Objectives | 3 | State specific objectives, including any prespecified hypotheses | Page 3: Introduction | | |
| **Methods** | | | | | |
| Study Design | 4 | Present key elements of study design early in the paper | Page 4-5: Methods | | |
| Setting | 5 | Describe the setting, locations, and relevant dates, including periods of recruitment, exposure, follow-up, and data collection | Page 4-5: Methods | | |
| Participants | 6 | *(a) Cohort study* - Give the eligibility criteria, and the sources and methods of selection of participants. Describe methods of follow-up<br><br>*Case-control study* - Give the eligibility criteria, and the sources and methods of case ascertainment and control selection. Give the rationale for the choice of cases and controls<br><br>*Cross-sectional study* - Give the eligibility criteria, and the sources and methods of selection of participants<br><br>*(b) Cohort study* - For matched studies, give matching criteria and number of exposed and unexposed | Page 4-5: Methods | RECORD 6.1: The methods of study population selection (such as codes or algorithms used to identify subjects) should be listed in detail. If this is not possible, an explanation should be provided.<br><br>RECORD 6.2: Any validation studies of the codes or algorithms used to select the population should be referenced. If validation was conducted for this study and not published elsewhere, detailed methods and results should be provided.<br><br>RECORD 6.3: If the study involved linkage of databases, consider use of a flow diagram or other graphical display to demonstrate the data linkage process, including the number of individuals with linked data at each stage. | Page 4-5: Methods |

| | | | | | |
|---|---|---|---|---|---|
| | | *Case-control study* - For matched studies, give matching criteria and the number of controls per case | | | |
| Variables | 7 | Clearly define all outcomes, exposures, predictors, potential confounders, and effect modifiers. Give diagnostic criteria, if applicable. | Page 4-5: Methods | RECORD 7.1: A complete list of codes and algorithms used to classify exposures, outcomes, confounders, and effect modifiers should be provided. If these cannot be reported, an explanation should be provided. | Supplementary appendix |
| Data sources/ measurement | 8 | For each variable of interest, give sources of data and details of methods of assessment (measurement).<br><br>Describe comparability of assessment methods if there is more than one group | Page 4-5: Methods | | |
| Bias | 9 | Describe any efforts to address potential sources of bias | Page 4-5: Methods | | |
| Study size | 10 | Explain how the study size was arrived at | Page 4-5: Methods, Page 6: Results | | |
| Quantitative variables | 11 | Explain how quantitative variables were handled in the analyses. If applicable, describe which groupings were chosen, and why | Page 4-5: Methods | | |
| Statistical methods | 12 | (a) Describe all statistical methods, including those used to control for confounding | Page 4-5: Methods | | |

| | | | | | |
|---|---|---|---|---|---|
| | | (b) Describe any methods used to examine subgroups and interactions<br><br>(c) Explain how missing data were addressed<br><br>(d) *Cohort study* - If applicable, explain how loss to follow-up was addressed<br><br>*Case-control study* - If applicable, explain how matching of cases and controls was addressed<br><br>*Cross-sectional study* - If applicable, describe analytical methods taking account of sampling strategy<br><br>(e) Describe any sensitivity analyses | | | |
| Data access and cleaning methods | | .. | | RECORD 12.1: Authors should describe the extent to which the investigators had access to the database population used to create the study population.<br><br>RECORD 12.2: Authors should provide information on the data cleaning methods used in the study. | Page 4-5: Methods |
| Linkage | | .. | | RECORD 12.3: State whether the study included person-level, institutional-level, or other data linkage across two or more databases. The methods of linkage and | Page 4-5: Methods |

| | | | | | |
|---|---|---|---|---|---|
| | | | | methods of linkage quality evaluation should be provided. | |
| **Results** | | | | | |
| Participants | 13 | (a) Report the numbers of individuals at each stage of the study (*e.g.*, numbers potentially eligible, examined for eligibility, confirmed eligible, included in the study, completing follow-up, and analysed)<br><br>(b) Give reasons for non-participation at each stage.<br><br>(c) Consider use of a flow diagram | Page 6: Results<br><br>Figure 1 | RECORD 13.1: Describe in detail the selection of the persons included in the study (*i.e.,* study population selection) including filtering based on data quality, data availability and linkage. The selection of included persons can be described in the text and/or by means of the study flow diagram. | Figure 1 |
| Descriptive data | 14 | (a) Give characteristics of study participants (*e.g.*, demographic, clinical, social) and information on exposures and potential confounders<br><br>(b) Indicate the number of participants with missing data for each variable of interest<br><br>(c) *Cohort study* - summarise follow-up time (*e.g.*, average and total amount) | Page 6: Results – demographic and clinical characteristics | | |
| Outcome data | 15 | *Cohort study* - Report numbers of outcome events or summary measures over time<br><br>*Case-control study* - Report numbers in each exposure | Page 6: Results – main analysis | | |

| | | | | | |
|---|---|---|---|---|---|
| | | category, or summary measures of exposure<br><br>*Cross-sectional study* - Report numbers of outcome events or summary measures | | | |
| Main results | 16 | (a) Give unadjusted estimates and, if applicable, confounder-adjusted estimates and their precision (e.g., 95% confidence interval). Make clear which confounders were adjusted for and why they were included<br><br>(b) Report category boundaries when continuous variables were categorized<br><br>(c) If relevant, consider translating estimates of relative risk into absolute risk for a meaningful time period | Not applicable | | |
| Other analyses | 17 | Report other analyses done—e.g., analyses of subgroups and interactions, and sensitivity analyses | Page 6-7: Results – secondary and sensitivity analyses | | |
| **Discussion** | | | | | |
| Key results | 18 | Summarise key results with reference to study objectives | Page 8: Discussion | | |
| Limitations | 19 | Discuss limitations of the study, taking into account sources of potential bias or imprecision. | Page 8-9: Discussion – Strengths and limitations | RECORD 19.1: Discuss the implications of using data that were not created or collected to answer the specific research question(s). Include discussion of | Page 8-9: Discussion – |

| | | | | | |
|---|---|---|---|---|---|
| | | Discuss both direction and magnitude of any potential bias | | misclassification bias, unmeasured confounding, missing data, and changing eligibility over time, as they pertain to the study being reported. | Strengths and limitations |
| Interpretation | 20 | Give a cautious overall interpretation of results considering objectives, limitations, multiplicity of analyses, results from similar studies, and other relevant evidence | Page 8: Discussion | | |
| Generalisability | 21 | Discuss the generalisability (external validity) of the study results | Page 8-9: Discussion – Strengths and limitations | | |
| **Other Information** | | | | | |
| Funding | 22 | Give the source of funding and the role of the funders for the present study and, if applicable, for the original study on which the present article is based | Page 12: Funding | | |
| Accessibility of protocol, raw data, and programming code | | .. | | RECORD 22.1: Authors should provide information on how to access any supplemental information such as the study protocol, raw data, or programming code. | |

*Reference: Benchimol EI, Smeeth L, Guttmann A, Harron K, Moher D, Petersen I, Sørensen HT, von Elm E, Langan SM, the RECORD Working Committee. The REporting of studies Conducted using Observational Routinely-collected health Data (RECORD) Statement. *PLoS Medicine* 2015; in press.

*Checklist is protected under Creative Commons Attribution (CC BY) license.

### Appendix S2: COVID-19 code lists

#### COVID-19 diagnostic codes

The codes used to identify COVID-19 diagnosis are listed below

| Code | Diagnostic description |
|---|---|
| **READ v2 Codes** | |
| 4J3R100 | 2019-nCoV (novel coronavirus) RNA detected |
| A795400 | Acute disease caused by SARS-CoV-2 |
| A795200 | COVID-19 confirmed by laboratory test |
| A795300 | COVID-19 confirmed using clinical diagnostic criteria |
| 38VO.00 | COVID-19 severity score |
| A7y0000 | Coronavirus as cause of dis classified to other chapters |
| A795.00 | Coronavirus infection |
| A795100 | Disease caused by 2019-nCoV (novel coronavirus) |
| F289.00 | Encephalopat due to SARS-CoV-2 |
| A076400 | Gastroenteri due to SARS-CoV-2 |
| G520800 | Myocarditis due to SARS-CoV-2 |
| A795500 | Ongoing symptomatic COVID-19 |
| F529.00 | Otitis media due to SARS-CoV-2 |
| H204.00 | Pneumonia due to SARS-CoV-2 |
| 43kB100 | SARS-CoV-2 antigen positive |
| H051100 | URTI due to SARS-CoV-2 |
| AyuDC00 | [X]Coronavirus infection, unspecified |
| AyuKL00 | [X]Coronavirus/cause/diseases classified to other chapters |
| **ICD-10** | |
| U07.1 | COVID-19, virus identified |
| U07.2 | COVID-19, virus not identified |
| **SNOMED Concept ID codes** | |
| 2537007017 | SARS-CoV |
| 2536663014 | SARS-CoV infection |
| 2832021000000118 | 2019-nCoV (novel coronavirus) antigen detection result positive |
| 2808081000000112 | 2019-nCoV (novel coronavirus) detected |
| 2837991000000116 | 2019-nCoV (novel coronavirus) detection result positive at the limit of detection |
| 2837121000000116 | 2019-nCoV (novel coronavirus) ribonucleic acid detected |
| 4361482017 | Acute COVID-19 |
| 2839411000000114 | Acute COVID-19 infection |
| 4361485015 | Acute disease caused by Severe acute respiratory syndrome coronavirus 2 |
| 2839421000000115 | Acute disease caused by severe acute respiratory syndrome coronavirus 2 infection |
| 3951313019 | Asymptomatic COVID-19 |
| 2838071000000117 | Asymptomatic SARS-CoV-2 (severe acute respiratory syndrome coronavirus 2) infection |
| 12802201000006117 | Confirmed 2019-nCoV (novel coronavirus) infection |
| 2826511000000115 | COVID-19 |

| | |
|---|---|
| 3947185011 | COVID-19 |
| 2826541000000119 | COVID-19 caused by SARS-CoV-2 (severe acute respiratory syndrome coronavirus 2) |
| 2799591000000111 | COVID-19 confirmed by laboratory test |
| 2799661000000118 | COVID-19 confirmed clinically |
| 2799621000000114 | COVID-19 confirmed using clinical diagnostic criteria |
| 3973484010 | COVID-19 detected |
| 2807871000000116 | Detection of 2019-nCoV (novel coronavirus) using polymerase chain reaction technique |
| 3973481019 | Detection of COVID-19 |
| 3973433014 | Detection of ribonucleic acid of 2019 novel coronavirus in nasopharyngeal swab |
| 3973463017 | Detection of ribonucleic acid of COVID-19 using polymerase chain reaction |
| 3973428010 | Detection of ribonucleic acid of Severe acute respiratory syndrome coronavirus 2 |
| 2838581000000112 | Detection of RNA (ribonucleic acid) of SARS-CoV-2 (severe acute respiratory syndrome coronavirus 2) in nasopharyngeal swab |
| 2838561000000115 | Detection of RNA (ribonucleic acid) of SARS-CoV-2 (severe acute respiratory syndrome coronavirus 2) in oropharyngeal swab |
| 2838541000000116 | Detection of RNA (ribonucleic acid) of SARS-CoV-2 (severe acute respiratory syndrome coronavirus 2) in sputum |
| 2838511000000117 | Detection of RNA (ribonucleic acid) of SARS-CoV-2 (severe acute respiratory syndrome coronavirus 2) using polymerase chain reaction |
| 2838481000000111 | Detection of SARS-CoV-2 (severe acute respiratory syndrome coronavirus 2) |
| 2838601000000115 | Detection of SARS-CoV-2 (severe acute respiratory syndrome coronavirus 2) antigen |
| 2826591000000112 | Detection of SARS-CoV-2 (severe acute respiratory syndrome coronavirus 2) using polymerase chain reaction technique |
| 3902357013 | Disease caused by 2019 novel coronavirus |
| 3902358015 | Disease caused by 2019-nCoV |
| 2808881000000118 | Disease caused by 2019-nCoV (novel coronavirus) |
| 3970750011 | Disease caused by Severe acute respiratory syndrome coronavirus 2 |
| 2808901000000115 | Disease caused by Wuhan 2019-nCoV (novel coronavirus) |
| 2832011000000112 | SARS-CoV-2 (severe acute respiratory syndrome coronavirus 2) antigen detection result positive |
| 2827041000000112 | SARS-CoV-2 (severe acute respiratory syndrome coronavirus 2) detected |
| 2831581000000117 | SARS-CoV-2 (severe acute respiratory syndrome coronavirus 2) RNA (ribonucleic acid) detection result positive |
| 2837101000000113 | SARS-CoV-2 (severe acute respiratory syndrome coronavirus 2) RNA (ribonucleic acid) detection result positive |
| 2837971000000115 | SARS-CoV-2 (severe acute respiratory syndrome coronavirus 2) RNA (ribonucleic acid) detection result positive at the limit of detection |
| 2837091000000117 | Severe acute respiratory syndrome coronavirus 2 ribonucleic acid detected |
| 2808101000000118 | Wuhan 2019-nCoV (novel coronavirus) detected |

| | |
|---|---|
| 2799651000000116 | Probable COVID-19 confirmed using clinical diagnostic criteria |
| 3970759012 | SARS (severe acute respiratory syndrome) coronavirus 2 RNA |
| 2833561000000113 | SARS-CoV-2 - severe acute respiratory syndrome coronavirus 2 |
| 2829511000000110 | SARS-CoV-2 (severe acute respiratory syndrome coronavirus 2) detection result positive |
| 2831751000000118 | SARS-CoV-2 (severe acute respiratory syndrome coronavirus 2) RNA (ribonucleic acid) qualitative existence in specimen |
| 3970626018 | Severe acute respiratory syndrome coronavirus 2 detected |

### Appendix S3: Propensity score matching plots

### The figure below shows the absolute standardised mean differences before and after matching

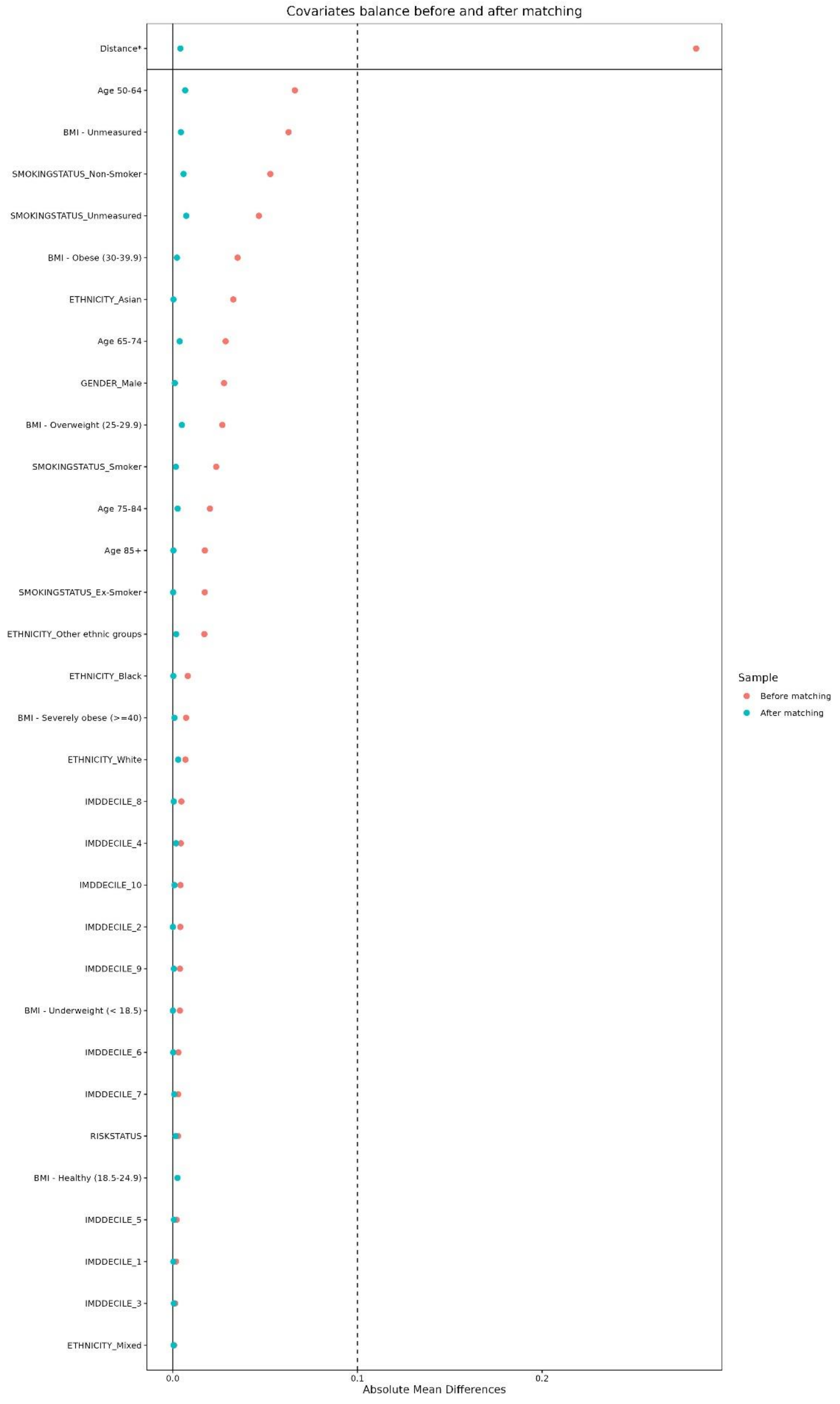

**The figure below shows the propensity score distribution before and after matching**

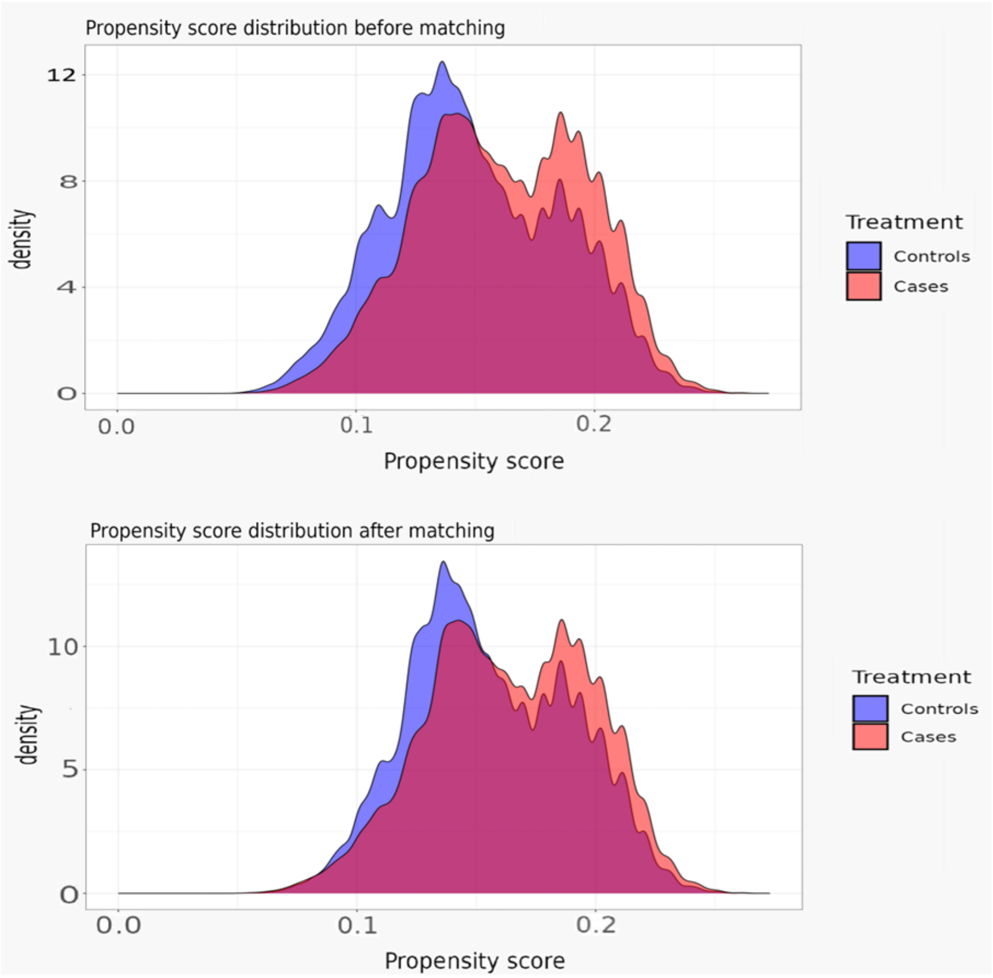

**Appendix S4: Social care outcome definitions**

**Key words used to identify social care use**

Social care resource use in the ‘Service Type Group’ and ‘Service Type Description’ fields was identified and classified into four pre-defined categories (care home, domiciliary care, respite care, and assessment). This classification was reported as binary yes/no variables, using automated code (regular expressions) to search for keywords, as per a previous study using the electronic social care records in Discover. There was no human review of the free text. The R packages ‘*tidyverse*’, ‘*stringr*’, and ‘*lubridate’* was used for string text searches and data manipulation. All strings were exact matches and searches were case insensitive.* The table below shows the keywords used to assign identified social care resource use to a category

| Social care type | Keyword(s) |
|---|---|
| Care Home | “*care home; nursing home; residential; accommodation; shared lives;”* |
| Domiciliary care | *“homecare; home care; domiciliary; nursing; day service; reablement; support at home; supported living; day care; extra care”* |
| Respite care | *“respite; residential short term”* |
| Assessment | *“assessment; review; plan”* |

**Summary of social care costs by setting**

Social care cost outcomes are described in the table below

| Variable | Cost per unit (2022/23) | Operational definition |
|---|---|---|
| Care home cost | £247 per day | Local authority own-provision residential care for older people (age 65+). Establishment cost plus personal living expenses and external services per permanent resident day. |
| Domiciliary care cost | £199.80 per day | Costs per weekday hour for a home care worker at £27 per hour with an assumption of 7.4 hours of domiciliary care provided per day. |
| Respite care cost | £68 per day | Local authority own-provision day care for older people (age 65+). £17 per client hour unit cost with an assumption of 4 hours of respite care provided per day. |
| Assessment cost | £49 per assessment | Needs assessments are conducted by a social care worker or an occupational therapist and last at least an hour. Social care assessments are assumed to last one hour each. Cost per hour will be estimated as the average of community |

| | | |
|---|---|---|
| | | occupational therapists and a social care worker (with and without qualification). Details below.<br><br>Community occupational therapist (local authority)<br><br>• £52 per hour with qualifications<br>• £48 per hour without qualification<br><br>Social worker (adult services)<br><br>• £53 per hour with qualifications<br><br>£44 per hour without qualification |

**Appendix S5: Demographic and clinical characteristic definitions and code lists**

Code lists used in this study can be accessed here [PLACEHOLDER – link to be added once accepted for publication]

**Definition of severe COVID-19 risk status**

Comorbidities indicating an increased risk of severe COVID-19 were identified using diagnosis codes, including READ v2, SNOMED CT codes in primary care, or ICD-10 codes in secondary care. These definitions were used to create a binary 'High-risk status' variable: individuals belonging to the clinical risk groups were classified as "High-risk," while those not belonging to these groups were classified as "Not high-risk.

Operational definitions for some comorbidities differ in this study compared to those in the Green Book as follows:

1) The definition of poorly controlled asthma will be based on the use of any oral corticosteroid in the 24 months prior to the date when comorbidity status is ascertained.
2) The chronic heart disease and vascular disease definitions used will be based on diagnosis codes and will not use the medication definition.
3) The risk group "Younger adults in long-stay nursing and residential care settings" is not included in this study given that the study population is restricted to individuals aged ≥50 years.
4) The risk group "Pregnancy" is not included in this study given that the study population is restricted to individuals aged ≥50 years.
5) Immunosuppression medications will not be included in the definition of immunosuppression because Discover only includes primary care medications, and not medications prescribed in hospital (such as biologic therapies used for systemic lupus erythematosus, rheumatoid arthritis, inflammatory bowel disease, scleroderma, and psoriasis)

**Definition of frailty**

The electronic and clinical frailty status closest to, and including, the index date, within two years of index date were derived from Read v2 codes that provide clinical or electronic frailty index records. All frailty-related entries were mapped to the following mutually exclusive frailty groups:

- Mild frailty (reference group)
- Moderate frailty
- Severe frailty
- Unknown

In the event that multiple frailty records existed for the same date, then the more severe frailty status was used. All patients were also assigned either: Record of frailty or No recorded frailty. The table below describes the code lists used to identify frailty.

| Code | Diagnostic description |
|---|---|
| **READ v2** | |
| 2Jd.. | Frailty |
| 2Jd0. | Mild frailty |
| 2Jd1. | Moderate frailty |
| 2Jd2. | Severe frailty |
| 69D9. | Frail elderly assessment |

| | |
|---|---|
| 38Q1. | OCAIRS |
| 38DW. | Canadian Study of Health and Ageing Clinical Frailty |
| 38GD. | Edmonton Frailty Scale |
| **ICD-10** | |
| R54 | Age-related physical debility |

**Definition of body mass index (BMI)**

BMI is recorded as ‘UNDERWEIGHT’(<18.5 kg/m$^2$), ‘HEALTHY’ (18.5–24.9 kg/m$^2$, ‘REFERENCE GROUP’), ‘OVERWEIGHT’(25–29.9 kg/m$^2$), ‘OBESE’(30–39.9 kg/m$^2$) and ‘SEVERELY OBESE’ (≥40 kg/m$^2$), and ‘UNKNOWN’. Where BMI was not recorded but a record for height and weight existed, BMI = weight (kg) ÷ height2 (meters) and categorised as above. Due to the infrequency in reporting of BMI, BMI will be derived within 2 years either side of (pseudo) index date, using the most recent point to and including the (pseudo) index date. The following table shows the codes used to identify BMI.

| CODE | Diagnostic description |
|---|---|
| **Read v2** | |
| 22K.. | Body Mass Index |
| 22K4. | BMI 25-29 – overweight |
| 22K5. | Body mass index 30+ – obesity |
| 22K6. | Body mass index less than 20 |
| 22K7. | BMI 40+ – severely obese |
| 22K8. | Body mass index 20-24 – normal |

**Definition of smoking status**

Smoking status is recorded as ‘SMOKER’, ‘EX-SMOKER’, ‘NON-SMOKER’, ‘UNKNOWN’. Due to the infrequency in reporting of smoking status, smoking status was derived within 2 years either side of (pseudo) index date, using the most recent point to and including the (pseudo) index date. The following table shows the codes used to identify smoking status.

| Code | Diagnostic description |
|---|---|
| **READ v2** | |
| 1371. | Never smoked tobacco |
| 1372. | Trivial smoker – < 1 cig/day |
| 1373. | Light smoker – 1-9 cigs/day |
| 1374. | Moderate smoker – 10-19 cigs/day |
| 1375. | Heavy smoker – 20-39 cigs/day |
| 1376. | Very heavy smoker – 40+ cigs/day |
| 1377. | Ex-trivial smoker (<1 / day) |
| 1378. | Ex-light smoker (1-9/day) |
| 1379. | Ex-moderate smoker (10-19/day) |
| 137A. | Ex-heavy smoker (20-39/day) |
| 137B. | Ex-very heavy smoker (40+/day) |
| 137b. | Ready to stop smoking |
| 137C. | Keeps trying to stop smoking |
| 137c. | Thinking about stopping smoking |
| 137d. | Not interested in stopping smoking |
| 137e. | Smoking restarted |
| 137F. | Ex-smoker – amount unknown |
| 137f. | Reason for restarting smoking |
| 137G. | Trying to give up smoking |
| 137h. | Min from wake to 1st tobacco consumption |
| 137H. | Pipe smoker |
| 137i. | Ex-tobacco chewer |
| 137I. | Passive smoker |
| 137I0 | Exposed to tobacco smoke – home |
| 137J. | Cigar smoker |

| | |
|---|---|
| 137j. | Ex-cigarette smoker |
| 137K. | Stopped smoking |
| 137K0 | Recently stopped smoking |
| 137L. | Current non-smoker |
| 137l. | Ex roll-up cigarette smoker |
| 137m. | Failed attempt to stop smoking |
| 137M. | Rolls own cigarettes |
| 137N. | Ex pipe smoker |
| 137n. | Total time smoked |
| 137O. | Ex cigar smoker |
| 137P. | Cigarette smoker |
| 137Q. | Smoking started |
| 137R. | Current smoker |
| 137S. | Ex smoker |
| 137T. | Date ceased smoking |
| 137U. | Not a passive smoker |
| 137V. | Smoking reduced |
| 137W. | Chews tobacco |
| 13p.. | Smoking cessation milestones |
| 9ko.. | Current smoker annual review - ESA |

**Definition of ethnicity**

Ethnicity was recorded in Discover on or before May 20, 2024. Participants were recorded as ‘WHITE’, ‘ASIAN OR ASIAN BRITISH’, ‘BLACK, BLACK BRITISH’, ‘MIXED’, ‘OTHER ETHNIC GROUPS’, ‘UNKNOWN’.

**Definition of geolocation**

Based on patient postcode, as recorded in Discover on May 20, 2024.

### Appendix S6: Secondary analysis

We compared social care use before and after COVID-19 infection to explore social care outcome among all individuals with COVID-19 including those not taken forward to the matched cohort. This establishes the pre-existing trends in social care use before COVID-19 infection

Social care events rate ratios and mean difference in costs for the secondary analysis. Overall and by subgroup.

**Rate Ratios with 95% CI**

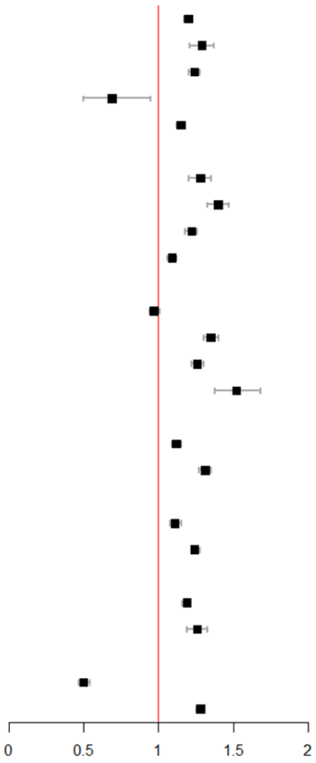


| Characteristics | RR | 95% CI | P-value |
|---|---|---|---|
| **Social care events** | | | |
| Overall social care events | 1.20 | (1.17, 1.22) | <0.0001 |
| Care home | 1.29 | (1.21, 1.37) | <0.0001 |
| Domiciliary care | 1.24 | (1.20, 1.28) | <0.0001 |
| Respite care events | 0.69 | (0.50, 0.95) | 0.0022 |
| Social care assessment | 1.15 | (1.12, 1.18) | 0.7163 |
| **Age at index** | | | |
| 50 - 64 | 1.28 | (1.20, 1.35) | 0.0107 |
| 65 - 74 | 1.40 | (1.33, 1.47) | <0.0001 |
| 75 - 84 | 1.22 | (1.18, 1.26) | <0.0001 |
| 85+ | 1.09 | (1.06, 1.12) | 0.0327 |
| **Calendar year at index** | | | |
| 2020 | 0.97 | (0.94, 1.01) | 0.3723 |
| 2021 | 1.35 | (1.30, 1.40) | <0.0001 |
| 2022 | 1.26 | (1.22, 1.30) | <0.0001 |
| 2023 | 1.52 | (1.38, 1.68) | 0.0055 |
| **Diagnosis Setting** | | | |
| Primary care | 1.12 | (1.09, 1.15) | <0.0001 |
| Secondary care | 1.31 | (1.27, 1.35) | <0.0001 |
| **Risk Group** | | | |
| High-risk of severe COVID | 1.11 | (1.08, 1.15) | 0.0117 |
| Not at high risk of severe COVID | 1.24 | (1.22, 1.28) | <0.0001 |
| **Frailty** | | | |
| Record of frailty | 1.19 | (1.16, 1.21) | <0.0001 |
| No recorded frailty | 1.26 | (1.19, 1.33) | 0.0047 |
| **Care home admission** | | | |
| Admitted prior to index | 0.50 | (0.47, 0.54) | <0.0001 |
| Not admitted prior to index | 1.28 | (1.25, 1.30) | <0.0001 |

**Mean difference in cost (£) with 95% CI**

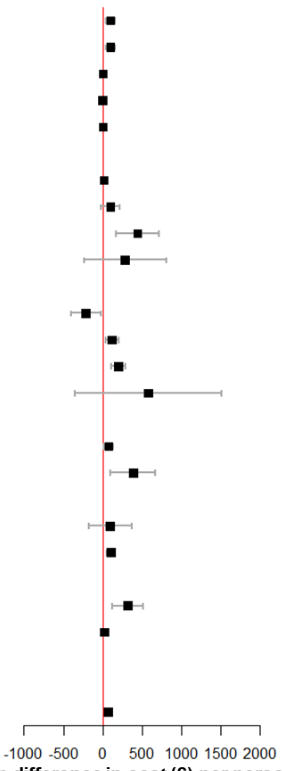


| Characteristics | Cost | 95% CI | P-value |
|---|---|---|---|
| **Social care events** | | | |
| Overall social care events | 99.74 | (39.13, 160.35) | <0.0001 |
| Care home | 96.67 | (36.14, 157.19) | <0.0001 |
| Domiciliary care | 2.72 | (-0.42, 5.86) | 0.0311 |
| Respite care events | -0.13 | (-0.27, 0.01) | 0.0016 |
| Social care assessment | 0.48 | (0.24, 0.72) | 0.6612 |
| **Age at index** | | | |
| 50 - 64 | 15.29 | (-22.59, 53.18) | 0.0160 |
| 65 - 74 | 97.86 | (-21.85, 217.58) | <0.0001 |
| 75 - 84 | 442.64 | (168.16, 717.12) | <0.0001 |
| 85+ | 283.66 | (-240.08, 807.40) | 0.0180 |
| **Calendar year at index** | | | |
| 2020 | -214.87 | (-404.12, -25.63) | 0.8620 |
| 2021 | 118.23 | (36.81, 199.65) | <0.0001 |
| 2022 | 195.44 | (106.86, 284.01) | <0.0001 |
| 2023 | 576.59 | (-357.56, 1510.75) | 0.0114 |
| **Diagnosis Setting** | | | |
| Primary care | 71.75 | (11.83, 131.66) | <0.0001 |
| Secondary care | 385.46 | (100.07, 670.86) | <0.0001 |
| **Risk Group** | | | |
| High-risk of severe COVID | 92.96 | (-179.45, 365.38) | 0.0676 |
| Not at high risk of severe COVID | 101.60 | (46.60, 156.60) | <0.0001 |
| **Frailty** | | | |
| Record of frailty | 318.89 | (126.90, 510.88) | <0.0001 |
| No recorded frailty | 17.81 | (-18.20, 53.81) | 0.0035 |
| **Care home admission** | | | |
| Admitted prior to index | 4134.63 | (1302.51, 6966.76)* | 0.1643 |
| Not admitted prior to index | 65.74 | (22.44, 109.04) | <0.0001 |

*Not plotted

**Appendix S7: Sensitivity analyses**

**Description of sensitivity analyses**

We utilised the PSM population created for the main analysis, excluding individuals that do not meet the relevant sensitivity analysis definitions. Valid sets (consisting of a COVID-19 case and at least one matched comparator that meet the relevant sensitivity analysis definitions) were retained. Individuals with COVID-19 without at least one matched comparator, or individual comparators not matched to a COVID-19 case were excluded from the relevant sensitivity analyses populations

| Sensitivity analyses | Description | Rationale |
|---|---|---|
| 1 | Individuals were censored at their second COVID-19 diagnosis. | Analysis varies the follow up period for social care use, testing whether results are consistent across different follow up periods. |
| 2 | SCRU and cost outcomes were assessed among individuals with no prior social care use | Analysis excludes pre-existing social care users to isolate new social care use that occurs post COVID-19 infection. Excludes individuals who had a high risk of social care, creating a healthier study population |
| 3 | SCRU and cost outcomes were assessed among individuals with no prior care home admission | Analysis isolates new care home admissions that occur post COVID-19 infection to determine whether the association is driven by a single care type, or if other types of social care are increased. |
| 4 | SCRU and cost outcomes were assessed among individuals with respiratory infections (any of pneumococcal disease, influenza, respiratory syncytial virus) in either primary or secondary care in the 6 months prior to index date | Analysis helps to clarify whether increased social care use is due to cumulative effects of multiple infections; assesses social care use in vulnerable populations |

Social care events rate ratios and mean difference in costs for the main analysis compared to the sensitivity analyses. Overall and by social care type.

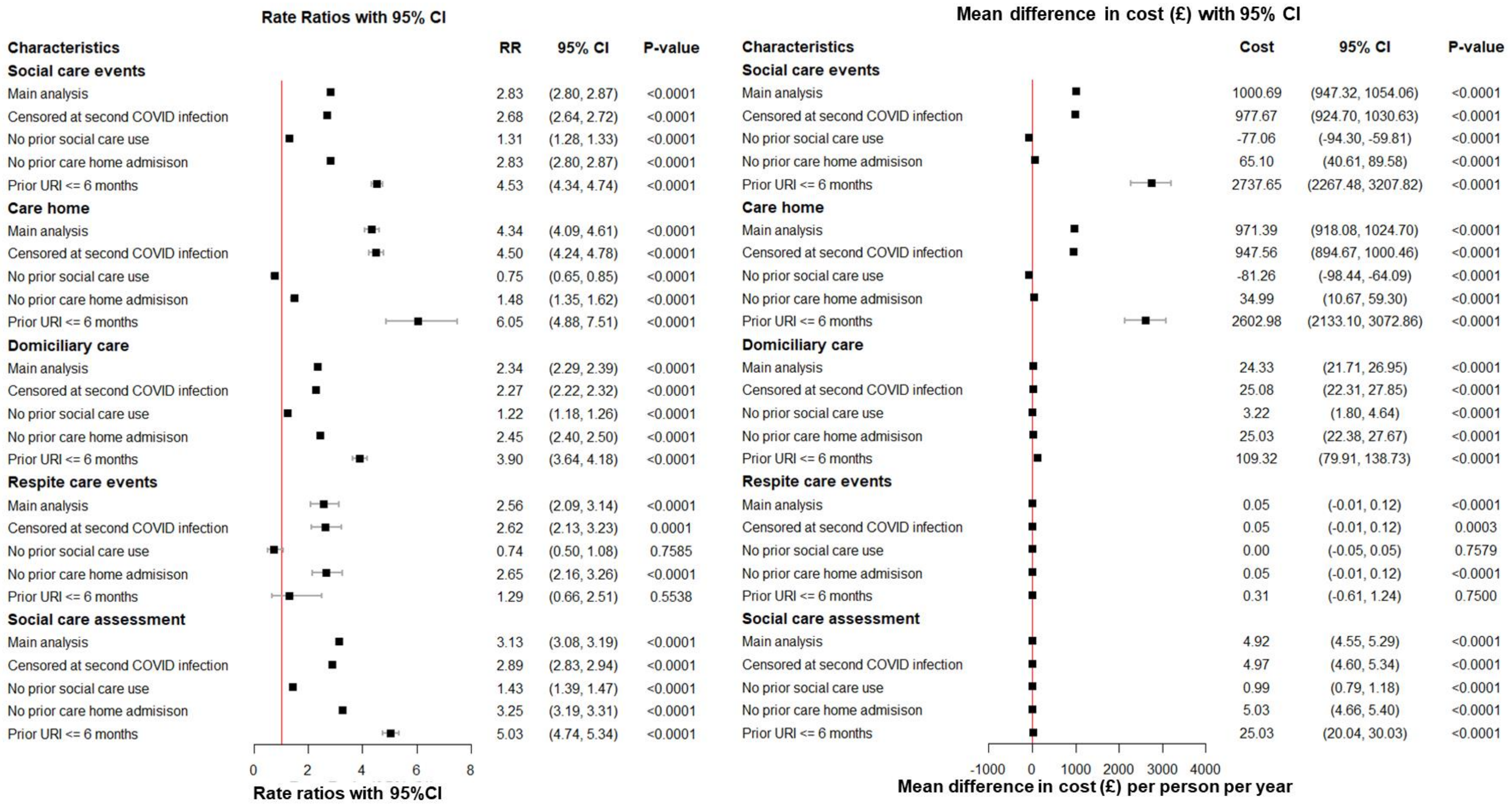

### Figure S2: Main analysis Kaplan-Meier Survival plot

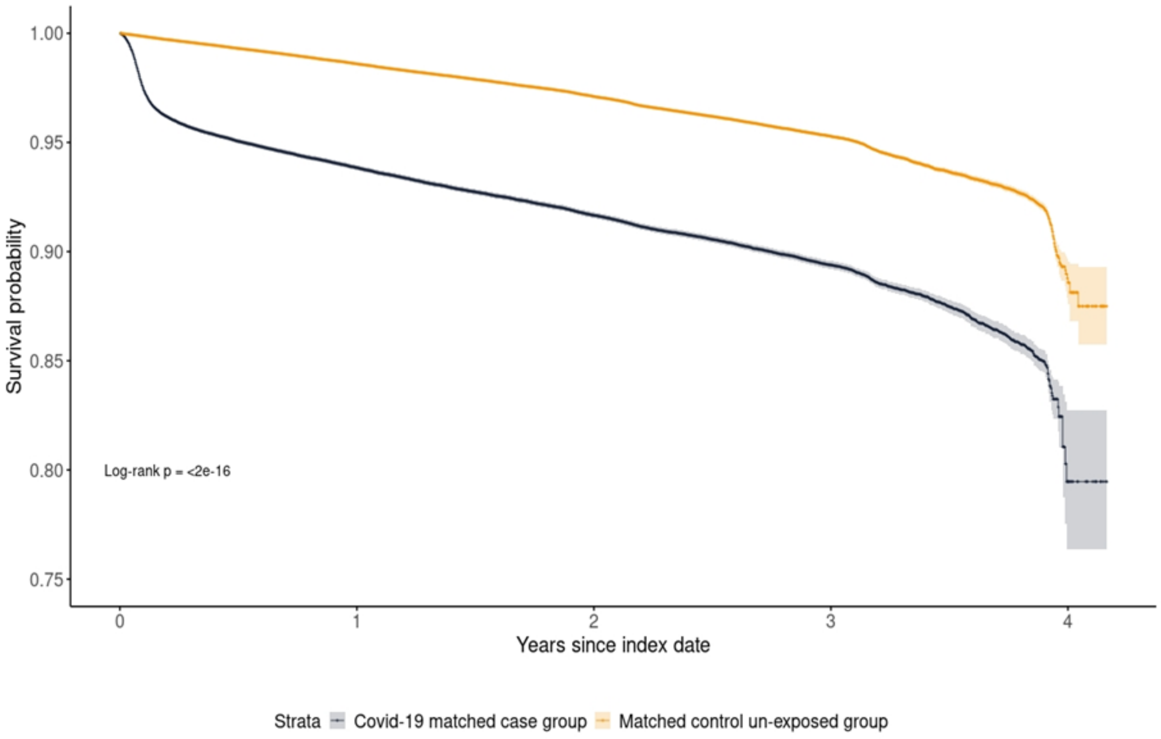
